# When the forest speaks again: in-depth genomic analysis of serotyping failure reveals geographic extension of sylvatic DENV-2 in southeastern Senegal, 2026

**DOI:** 10.64898/2026.10.19.26365950

**Authors:** Idrissa Dieng, Agathe Shella Efire, Samba Niang Sagne, Marie Henriette Dior Ndione, Diogop Camara, Serge Freddy Moukaha Doukanda, Ndongo Dia, Boubacar Diallo, Mamadou Aliou Barry, Gamou Fall, Oumar Faye

## Abstract

In August 2026, eight febrile patients in the Kédougou and Tambacounda regions of Senegal tested positive by pan-flavivirus and pan-dengue RT-PCR but could not be assigned to a serotype. In-depth genome sequencing identified sylvatic dengue virus serotype 2 (DENV-2, genotype VI). These findings indicate the re-emergence and geographic spread of DENV-2/GVI beyond its historical focus.

## The Study

As part of the ongoing syndromic surveillance program for febrile illness in Senegal (the 4S sentinel network), eight patients with acute febrile illness in the Kédougou (n = 6) and Tambacounda (n = 2) regions of southeastern Senegal were confirmed as dengue virus–infected between 13 August and 17 September 2026. All eight samples were positive for dengue RNA by RT-qPCR using a previously described protocol (1). Cycle threshold (Ct) values ranged from 26.6 to 35.12, confirming the presence of DENV RNA at a detectable viral load. Unexpectedly, none of the samples could be assigned to a serotype with the CDC dengue typing kit, pointing to a potential assay target failure as previously described (2,3). This pattern suggested circulation of sylvatic DENV-2 (genotype VI, DENV-2/GVI), consistent with earlier reports (4,14).

Globally, DENV-2/GVI circulation has been documented in several West African countries Nigeria, Senegal, Côte d’Ivoire, Guinea, and Guinea-Bissau and in Malaysia (Southeast Asia) (7,8). Some of these studies reported human involvement in the transmission chain (Figure 1, panel A). Two recent studies described contemporary DENV-2/GVI circulation in Senegal in 2020–2021 (4) and in Malaysia in 2024 (6). In Senegal, the Kédougou region is the recognized focus of DENV-2/GVI emergence, where virus circulation has been documented since 1970 (5).

**Figure 1.**
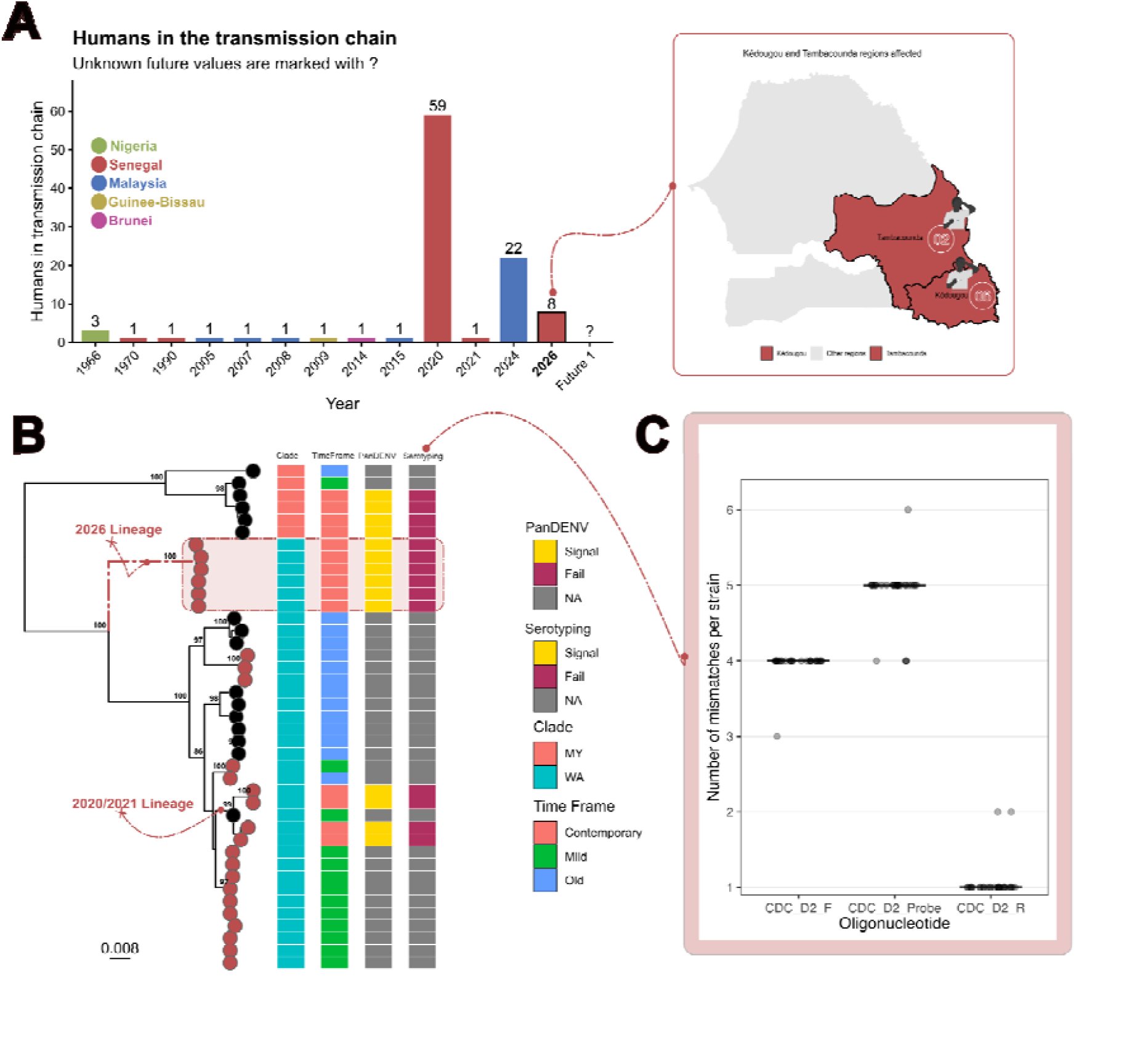
Emergence and genomic characterization of a 2026 sylvatic DENV-2 (DENV-2/GVI) lineage in southeastern Senegal. (A) Number of humans in the documented transmission chain per year, colored by country of origin (Nigeria, Senegal, Malaysia, Guinea-Bissau, Brunei). Counts rose from sporadic single cases (1968–2015) to larger clusters in 2020 (n = 59), 2024 (n = 22) and the emerging 2026 outbreak (n = 8); future values remain unknown (marked “?”). The inset map shows the affected Kédougou and Tambacounda regions of southeastern Senegal. (B) Maximum-likelihood phylogeny of DENV-2 genomes with ultrafast bootstrap support values shown at nodes. Tip symbols denote sequence source (brown = Senegalese/contemporary strains; black = reference). Annotation columns indicate assigned clade (MY = Cosmopolitan/Malaysia-associated, WA = West Africa), sampling time frame (Contemporary, Mild, Old), and diagnostic performance for the PanDENV and serotype-specific (Serotyping) assays (Signal = detected, Fail = not detected, NA = not tested). The highlighted 2026 lineage falls within the WA clade and is distinct from the earlier 2020/2021 lineage. Scale bar, 0.008 nucleotide substitutions per site. (C) Number of nucleotide mismatches per strain between the 2026-lineage genomes and each oligonucleotide of the CDC DENV-2 RT-PCR assay (forward primer “CDC D2 F”, probe “CDC D2 Probe”, reverse primer “CDC D2 R”). Mismatches concentrate in the forward primer (3–4) and probe (2–5) binding regions, consistent with the serotyping assay failures seen in panel B, whereas the reverse primer remains largely conserved (1 mismatch).

Following the workflow previously described by Dieng and colleagues (14), we performed in-depth genomic characterization of the detected dengue strains (n = 8). BLAST and phylogenetic analyses assigned all successfully recovered sequences (n = 6) to the sylvatic DENV-2 genotype (DENV-2/GVI). These sequences formed a lineage distinct from the strains obtained in Kédougou in 2020, falling within the historical West African sylvatic clade (Figure 1, panel B).

Kédougou, in the Sudano-Guinean zone of southeastern Senegal, holds the only documented sylvatic dengue focus in West Africa. DENV-2 was first isolated there in 1970, and entomologic surveillance since then has documented periodic amplifications of the sylvatic cycle among canopy *Aedes* mosquitoes (*Ae. furcifer, Ae. taylori, Ae. luteocephalus*) and non-human primates, separated by silent intervals of 5–8 years and uncorrelated with rainfall (5). The most recent documented amplification in Senegal caused 59 confirmed human cases between November 2020 and February 2021 (4) (Figure 1, panel A). In contrast, the Tambacounda cases lie outside the area in which sylvatic DENV-2 human infections have previously been documented. This suggests possible under-ascertainment in this region, a recent spillover, or travel-associated spread between the two regions.

We aligned the six 2026 genomes against the binding sites of the serotype-specific assay (Figure 1, panel C). The serotyping failure is explained by primer and probe mismatches between the CDC dengue typing kit and the newly detected sylvatic lineage. Across all analyzed DENV-2/GVI sequences (previously available sequences and those newly characterized here), the DENV-2 forward primer carried 3–4 mismatches, the reverse primer 1, and the probe 2–5, including mismatches within the terminal 3 nucleotides of the 3′ end, where mismatches most strongly depress amplification efficiency (11). Fewer mismatches were present in the 2020 Kédougou sylvatic genomes and in the shared historical West African sylvatic sequences, and none were present in the cosmopolitan-genotype sequences on which the assay was validated.

In contrast, the pan-dengue and pan-flavivirus assays target conserved regions (the 3′ UTR and NS5, respectively) (1,10), consistent with their retained sensitivity (Table 1).

**Table 1.** RT-qPCR assays applied to the DENV-2/GVI–positive samples.

| Assay | Target region | Result |
| --- | --- | --- |
| Pan-dengue real-time RT-PCR (1) | 3' UTR | Positive, 8/8 |
| DENV serotype-specific real-time RT-PCR (2) | E gene | No amplification, 0/8 |
| Pan-flavivirus RT-PCR (10) | NS5 | Positive, 8/8 |

The operational consequence is that an algorithm that screens with a pan-dengue assay and then serotypes will flag these infections as dengue but leave them unclassified, whereas an algorithm that screens with serotype-specific assays alone as used in several laboratories in the region will record them as negative (3). Sylvatic DENV-2 circulation in Kédougou is therefore likely under-ascertained, and the six cases reported here should be read as a minimum estimate.

DENV-2/GVI re-emerged in Kédougou in 2026, six years after the 2020 amplification, consistent with the 5-to-8-year periodicity described across five decades of surveillance (5,12). This interval argues that the sylvatic cycle in southeastern Senegal is maintained continuously in canopy *Aedes* and non-human primates, with human cases marking amplification peaks rather than new introductions. The contribution of amplifying small mammals should also not be excluded (13).

Two findings extend what was reported in 2020. First, human sylvatic DENV-2 cases occurred in the Tambacounda region, outside the Kédougou focus where most previously documented human infections and all entomologic amplifications have been recorded (5). Whether this reflects expansion of the sylvatic cycle itself, movement of infected people out of the focus, or surveillance that has only now looked in the right place, it enlarges the area in which sylvatic dengue must be considered in a febrile patient. Second, the strains now circulating are invisible to a serotyping assay in routine use. The 2020 study concluded that sylvatic strains might not require adaptation to enter urban transmission cycles and called for updated molecular diagnostic tools (4); the present cases show that this call was warranted.

The public health implications are concrete. Serotype is used to interpret dengue epidemiology, to anticipate severe disease arising from secondary infection with a heterologous serotype, and to guide vaccine policy; an infection recorded as dengue of unknown serotype contributes to none of these. Where serotype-specific assays are the primary screen, such infections are missed entirely. We recommend that laboratories in the region retain a pan-dengue or pan-flavivirus screening step upstream of serotyping, that serotype-specific assays be redesigned or supplemented with a sylvatic DENV-2–specific primer set such as the one used here, and that any dengue-positive, serotype-indeterminate sample from the Kédougou region be sequenced.

The claim of geographic extension requires caution: the Kédougou region was created from the Tambacounda region in 2008, so comparing present and historical administrative units can produce an apparent extension where none exists. Sylvatic dengue cannot be eliminated, because its reservoir lies in wildlife; detecting it depends entirely on assays that can see it, supported by an extensive One Health strategy monitoring the virus in humans, vectors, reservoirs, and the environment.

## Data Availability

All data produced in the present study are available upon reasonable request to the authors

## Acknowledgments

We thank the clinical and laboratory staff of the Virology Department at Institut Pasteur de Dakar, the field team, and the 4S network team.

## Data availability

Data are available upon request

## Author biography

Dr. Idrissa Dieng is a research scientist at Institut Pasteur de Dakar, Dakar, Senegal. His research interests include the surveillance, diagnosis, and molecular evolution of arboviruses, with a primary focus on dengue virus.

## Notes

### Competing Interest Statement

The authors have declared no competing interest.

### Author Declarations

All samples were deidentified.

## References

1. Wagner D, de With K, Huzly D, Hufert F, Weidmann M, Breisinger S, Eppinger S, Kern WV, Bauer TM. Nosocomial acquisition of dengue. Emerg Infect Dis. 2004 Oct;10(10):1872–3. doi: 10.3201/eid1010.031037. PMID: 15504282; PMCID: PMC3323263.

2. Santiago GA, Vergne E, Quiles Y, Cosme J, Vazquez J, Medina JF, et al. Analytical and clinical performance of the CDC real time RT-PCR assay for detection and typing of dengue virus. PLoS Negl Trop Dis. 2013;7(7):e2311.

3. Dieng I. Systematic misclassification of sylvatic dengue virus 2 (DENV-2) infections as “undetermined serotype”: implications in routine RT-qPCR surveillance. Trop Med Health. 2025;53. doi:10.1186/s41182-025-00827-0.

4. Dieng I, Diarra M, Sadio BD, Sow B, Gaye A, Diallo A, Faye M, Ndione MHD, Diallo D, Sankhe S, Ndiaye M, Danfakha F, Diop B, Sall AA, Fall G, Faye O, Loucoubar C, Faye O, Weaver SC, Diallo M, Barry MA, Diagne MM. Reemergence of Sylvatic Dengue Virus Serotype 2 in Kedougou, Senegal, 2020. Emerg Infect Dis. 2024 Apr;30(4):770–774. doi: 10.3201/eid3004.231301. PMID: 38526209; PMCID: PMC10977847.

5. Diallo M, Ba Y, Sall AA, Diop OM, Ndione JA, Mondo M, et al. Amplification of the sylvatic cycle of dengue virus type 2, Senegal, 1999–2000: entomologic findings and epidemiologic considerations. Emerg Infect Dis. 2003;9(3):362–7.

6. Suppiah J, Zulkifli MMS, Adiee AH, Zainudin NAZ, Dukut Soeharto ME, Ramli N, Wan MK, Sulong MR, Suli Z, Mohd Zain R. Genomic evidence links human dengue cases with undetermined serotypes to sylvatic lineages. Trop Med Health. 2025 Aug 25;53(1):114. doi: 10.1186/s41182-025-00795-5. PMID: 40851063; PMCID: PMC12376745.

7. Franco L, Di Caro A, Carletti F, Vapalahti O, Renaudat C, Zeller H, et al. First report of sylvatic DENV-2–associated dengue haemorrhagic fever in West Africa. PLoS Negl Trop Dis. 2011;5(8):e1251.

8. Vasilakis N, Shell EJ, Fokam EB, Mason PW, Hanley KA, Estes DM, et al. Evolutionary processes among sylvatic dengue type 2 viruses. J Virol. 2007;81(17):9591–5.

9. Dieng I, Cunha M, Diagne MM, Sembène PM, Zanotto PMA, Faye O, Faye O, Sall AA. Origin and Spread of the Dengue Virus Type 1, Genotype V in Senegal, 2015-2019. Viruses. 2021 Jan 4;13(1):57. doi: 10.3390/v13010057. PMID: 33406660; PMCID: PMC7824722.

10. Patel P, Landt O, Kaiser M, Faye O, Koppe T, Lass U, Sall AA, Niedrig M. Development of one-step quantitative reverse transcription PCR for the rapid detection of flaviviruses. Virol J. 2013 Feb 14;10:58. doi: 10.1186/1743-422X-10-58. PMID: 23410000; PMCID: PMC3616844.

11. Stadhouders R, Pas SD, Anber J, Voermans J, Mes TH, Schutten M. The effect of primer-template mismatches on the detection and quantification of nucleic acids using the 5′ nuclease assay. J Mol Diagn. 2010;12(1):109–17.

12. Vasilakis N, Cardosa J, Hanley KA, Holmes EC, Weaver SC. Fever from the forest: prospects for the continued emergence of sylvatic dengue virus and its impact on public health. Nat Rev Microbiol. 2011;9(7):532–41.

13. Cinkovich SC, Althouse BM, Hitchings MDT, Bonnaire-Fils P, Diop OM, Faye O, Faye EHA, Diallo D, Sadio BD, Sow A, Faye O, Diallo M, Benefit B, Watts DM, Sall AA, Weaver SC, Hanley KA, Cummings DAT. Seroprevalence of dengue virus antibodies among multiple species of non-human primates in Senegal suggests that sylvatic dengue virus is maintained in non-primate reservoirs in this region. PLoS Negl Trop Dis. 2026 Jan 27;20(1):e0013946. doi: 10.1371/journal.pntd.0013946. PMID: 41592115; PMCID: PMC12863672.

14. Dieng I, Sagne SN, Ndiaye M, Barry MA, Talla C, Mhamadi M, Balde D, Toure CT, Diop B, Sall AA, Fall G, Loucoubar C, Faye O and Faye O (2022) Detection of human case of dengue virus 2 belonging to sylvatic genotype during routine surveillance of fever in Senegal, Kolda 2021. Front. Virol. 2:1050880. doi: 10.3389/fviro.2022.1050880

